# SONIVA: Speech recOgNItion Validation in Aphasia

**DOI:** 10.1101/2025.06.03.25328889

**Authors:** Giulia Sanguedolce, Cathy J. Price, Sophie Brook, Dragos C. Gruia, Niamh V. Parkinson, Patrick A. Naylor, Fatemeh Geranmayeh

## Abstract

Post-stroke aphasia is a major contributor to language impairment and neuro-disability worldwide, making automated assessment a critical research priority. However, the development of clinically validated automatic speech recognition (ASR) systems remains limited by the lack of large, annotated datasets that capture aphasia’s heterogeneous and unpredictable manifestations. We introduce SONIVA (Speech recOgNItion Validation in Aphasia), the largest and most richly annotated database of pathological speech to date, comprising recordings from ≈1,000 stroke survivors (including over 200 longitudinally) and ≈7,000 age-matched controls. Current annotations include 576 patients (mean age: 61.23 ± 13.23 years; 69.81% male) and 104 controls (mean age: 61.05 ± 12.05 years; 34% male), with rich linguistic coding, orthographic and international phonetic alphabet transcriptions. Foundation models finetuned on SONIVA extract linguistic features that correlate with expert transcriptions (Spearman’s r = 0.86 - 0.79; p *<* 0.0001), while acoustic classifiers achieve 93% stroke classification accuracy. These results position SONIVA as a critical resource that can transform rehabilitation through objective, scalable speech assessment.

---

Acquired speech and language disorders pose a significant challenge in neurological diseases, affecting millions of individuals worldwide. These impairments arise from a wide range of conditions, including stroke, traumatic brain injury (TBI), and neurodegenerative diseases such as dementia and Parkinson’s disease [1]. Speech disorders are particularly disabling because they disrupt communication, social interaction, and functional independence, leading to increased healthcare needs, prolonged hospital stays, and higher mortality and morbidity rates. Stroke is one of the leading causes of acquired speech and language disorders, with up to 40% of survivors experiencing communication impairments, ranging from motor speech disorders (dysarthria, apraxia of speech) to linguistic deficits (aphasia). Among these conditions, aphasia—a language disorder affecting speech production, comprehension, reading, and writing—is particularly prevalent in stroke, affecting one-third of survivors, though it also occurs in dementia and TBI [1]. Aphasia alone leads to significant healthcare burdens, with longer hospitalisation times and increased reliance on rehabilitation and caregiving services. The economic burden of post-stroke aphasia is estimated at 3.41 billion USD annually, including medical costs, assistive technologies, lost productivity, and caregiver support [2]. Given these substantial clinical and socioeconomic impacts, improving the diagnosis and treatment of aphasia and other communication disorders has been recognised as a critical research priority for patients and stakeholders [3]. A major limitation of current clinical practice is the lack of scalable, objective tools for speech assessment. In this context, artificial intelligence (AI)-powered speech recognition technology presents a promising solution. By leveraging automated speech recognition (ASR) models trained on aphasic speech cohorts, AI can facilitate earlier diagnosis, continuous monitoring, and more accessible rehabilitation, which could be game-changing for both assessment and long-term management.

Datasets from stroke-induced speech impairments represent a particularly strong candidate for training ASR models, not only due to the high prevalence of aphasia but also because post-stroke deficits exhibit significant heterogeneity both across patients and within the same individual over time [4]. Unlike neurodegenerative diseases, where speech deterioration is progressive and relatively predictable, stroke-related impairments emerge acutely, with highly variable severity and recovery trajectories depending on the location and extent of brain damage. This variability presents both a challenge and an opportunity for developing robust pathological speech models capable of handling diverse speech deficits, applicable to a multitude of neurological disorders [5].

Stroke survivors typically experience a range of linguistic and motor impairments that collectively compromise communication and daily functioning. Among these, aphasia primarily affects the linguistic system [6], characterised by phonemic errors (incorrect sounds), semantic paraphasias (meaning-related word substitutions), disrupted speech rate, and various dysfluencies [7]. Patients with aphasia may also exhibit syntactic errors (omitting function words and grammatical morphemes), anomia (word-finding difficulties), and neologisms (invented words) [8].

In addition to the linguistic symptoms above, motor speech impairments are another major challenge for stroke survivors, arising from damage to the control of oro-facial, respiratory, and articulatory muscles. These impairments manifest as hypophonia (reduced volume), a strained or hoarse voice, breathy speech with shortened phrase outputs, and gasping between utterances [9]. Additionally, stroke can lead to dysarthria (slurred speech) and apraxia of speech (disruptions in motor planning and sequencing of sounds) [10], further complicating verbal communication.

Beyond language and motor deficits, post-stroke recovery is often further hindered by cognitive impairments, which frequently co-occur with aphasia and motor speech disorders [11]–[15]. Deficits in attention, memory, problem-solving, and executive functioning—all essential components for effective communication—often accompany aphasia in patients with stroke. The combined impact of cognitive and communication impairments extends beyond the clinical setting, disrupting social interactions and diminishing quality of life [16]. This interplay between language and cognition complicates both assessment and intervention, underscoring the need for comprehensive approaches to the management of post-stroke communication disorders.

Considering the intricate and diverse nature of post-stroke language impairments, it is no surprise that diagnosing and treating post-stroke aphasia remains a challenge. With respect to rehabilitation, the evidence supports early [17] and intensive [18] speech therapy, which is difficult to administer in most healthcare settings. While meeting these intensive therapy requirements poses significant challenges for healthcare providers and has broad social and public health implications [19], technological innovations offer promising solutions. ASR systems trained on comprehensive pathological speech databases can potentially facilitate not only a more efficient diagnosis but also frequent and remote therapy sessions, making rehabilitation more accessible to patients with mobility limitations or those in low-income or remote settings [20].

Despite recent advances in ASR applications for healthy speech, their performance remains inadequate for clinical applications in pathological speech, with word error rates (WER) ranging from 21% to 45% [21]–[23] compared to 9%-13% on healthy speech [24]. The primary obstacle is the lack of large and annotated databases that capture the diverse and complex impairment patterns alluded to earlier [25]–[27]. Significant progress has been made in developing pathological speech databases, contributing to both linguistic research and ASR advancements. Nevertheless, these resources have some limitations.

AphasiaBank [28], one of the pioneers in the field, and part of a larger speech database (TalkBank), contains data from over 400 people with all-cause aphasia and 200 control participants, featuring discourse tasks including picture descriptions, story retelling, and procedural discourse. Recent studies leveraging AphasiaBank have demonstrated significant progress in ASR applications using architectures like MT-DNN with BLSTM (Multi-Task Deep Neural Network with Bidirectional Long Short-Term Memory) [29], advanced attention mechanisms such as E-Branchformer and Conformer models [30] or Whisper [31]. However, AphasiaBank presents some limitations, including the absence of i) comprehensive clinical, functional and cognitive outcome data; ii) longitudinal speech data to investigate the temporal trajectory of change in speech within individuals; and iii) neuroanatomical data to facilitate lesion-language relationships.

Smaller databases focusing on specific motor speech impairments include the UA-Speech (N=15) [32] and the TORGO (N=8) [33] databases. Beyond their small sample size, these databases are constrained by the limited scope on short-form speech (e.g., single words or short phrases), which further confines their general applicability for ASR development. In such cases, the ASR may be learning features of the recording environment rather than actual speech impairment patterns [34]. Longer-form speech databases dedicated to dysfluent speech, such as SEP-28K database [35], provide audio clips labelled with stuttering types but lack word-by-word transcriptions, limiting their utility for supervised ASR training.

Therefore, current pathological speech databases remain inadequate for training ASR models tailored to aphasic speech. While datasets such as AphasiaBank, UA-Speech, and TORGO have significantly contributed to speech and language research, they suffer from critical limitations, including small sample sizes, lack of word-by-word transcriptions, inconsistencies in speech tasks, and missing clinical, functional, and neuroanatomical data. Additionally, the absence of longitudinal recordings limits the ability to study how speech impairments evolve over time, restricting their applicability for developing robust ASR models.

To overcome these limitations, a comprehensive pathological speech database should not only provide a larger and more diverse speaker pool but also ensure the inclusion of both long-form and short-form speech samples to capture a broad spectrum of impairments. Standardised, high-quality annotations detailing phonetic distortions, semantic errors, and dysfluencies are essential to enhance ASR training and evaluation. Additionally, integrating clinical and neuroanatomical data would allow for a deeper understanding of the relationship between speech deficits and brain damage, while longitudinal recordings would facilitate the study of recovery trajectories over time.

Here, we introduce SONIVA (Speech recOgNItion Validation in Aphasia), a large-scale database specifically designed to address these limitations. SONIVA comprises ≈1000 speech samples from stroke survivors, featuring high-quality recordings, meticulous linguistic annotations, and paired clinical data. Beyond database construction, we demonstrate how ASR models fine-tuned on a subset of SONIVA can extract clinically meaningful linguistic features and how acoustic features from SONIVA can be leveraged for disease classification.

## I. METHODS

### A. The SONIVA Database

The SONIVA database is composed of speech from stroke survivors and control participants. The database includes speech recordings of two long-form picture description tasks and three short-form single-word tasks (naming, reading, and repetition). The short-form tasks originate from participants in the Imperial Comprehensive Cognitive Assessment in Cerebrovascular Disease (IC3) study [36], [37], whilst both IC3 and Predicting Language Outcome and Recovery after Stroke (PLORAS) [38] study participants contributed to the longform picture description tasks. Both studies received ethical approval from the UK Health Research Authority^1^, and all participants provided written informed consent.

IC3 speech data were collected from consecutive patients with clinically confirmed acute stroke, recruited from Imperial College Healthcare NHS Trust hospitals in England. Patients completed cognition and language tests using the IC3 online assessment battery [36], [37] immediately after stroke and at 3, 6, and 12 months post-stroke. Details of all tasks, including language tasks, have been described elsewhere [36]. The assessment typically lasted 90-120 minutes, with duration varying based on impairment severity, with an unlimited number of breaks given to patients where it was clinically indicated. Additional comprehensive clinical data including Montreal Cognitive Assessment (MoCA) [39], functional outcomes with the Modified Rankin Scale(mRS) [40] and the stroke severity with the National Institutes of Health Stroke Scale (NIHSS) [41], as well as neuroimaging MRI scanning, all captured longitudinally after stroke, were acquired as detailed in the study protocol [36], [37]. Speech and cognitive data were also collected from 7,000 UK adults aged over 40 without neurological conditions, serving as a control sample via the online IC3 assessment tool hosted on the Cognitron platform^2^. PLORAS speech data were collected from stroke survivors recruited across multiple UK sites via the NIHR Clinical Research Network. The speech data were supplemented with demographic details (age, gender, handedness, and time poststroke), manual scoring of spontaneous speech, and gross anatomical information on lesion location [38].

SONIVA integrates language assessment protocols from both PLORAS and IC3, each employing distinct methodological approaches to language evaluation. Among the 22 tasks in IC3’s assessment battery, six focus on language, with four specifically evaluating spoken output (Table I). Both IC3 and PLORAS incorporate picture description tasks from the Comprehensive Aphasia Test (CAT) [42], with IC3 additionally using a custom-designed beach scene. These picture description tasks elicit complete utterances (e.g., “there is a cat”) rather than single-word responses (“cat”), enabling assessment of grammatical structures and descriptive capabilities. Additionally, IC3 uniquely includes single-word tasks to evaluate spoken fluency, self-correction, and phonological processing, integrating semantic assessment within the naming components.

**TABLE I.** IC3 Language Assessment Tasks

| Task | Purpose | Method | Trials |
| --- | --- | --- | --- |
| Picture Description | Assess spontaneous speech production and fluency | Describe two black-and-white line drawings in complete sentences | 2 images, 90 seconds each |
| Naming | Assess semantic object recognition and verbal output | Name line drawings from related to item from Boston Naming Test | 30 items |
| Reading | Evaluate visual orthographic analysis and verbal output | Read aloud written words on screen | 11 items (including 3 non-words) |
| Repetition | Test auditory phonological analysis and verbal output | Repeat spoken words | 20 items (from 40-word pool) |

**TABLE II.** Summary of training and test splits for acoustic feature classification, separated by patients and non-stroke controls.

| Dataset | Mean Age (SD) | Male (%) | Duration (min) | File Count |
| --- | --- | --- | --- | --- |
| **Train set** |  |  |  |  |
| *Patients* | 61.20 (13.78) | 71.21 | 767.16 | 603 |
| *Controls* | 61.99 (9.46) | 52.21 | 112.59 | 133 |
| **Total** | **61.34 (12.98)** | **68.63** | **879.75** | **736** |
| **Test set** |  |  |  |  |
| *Patients* | 59.43 (11.74) | 67.95 | 90.78 | 65 |
| *Controls* | 67.00 (9.04) | 54.55 | 18.28 | 24 |
| **Total** | **61.47 (11.56)** | **65.00** | **109.06** | **89** |

Note: Mean age is represented in years and file count represents the number of unique speech files, with multiple files per patient.

### B. Speech Annotation Methodology

The annotation process involved two distinct protocols summarised in Table I, divided into picture description and single-word tasks (Naming, Reading and Repetition):

#### Picture Description Annotation

Audio and video recordings were manually transcribed using the Computerised Language ANalysis (CLAN) program [43], following the Codes for the Human Analysis of Transcripts (CHAT) format [44]. Transcription was conducted by researchers trained in speech and language therapy. The annotation captured five key dimensions:

##### Broad Transcription

Each participant’s utterance (“PAR”) was transcribed orthographically with a corresponding IPA (International Phonetic Alphabet) line (“%pho”), capturing both word-level and phonetic-level information. This feature makes SONIVA unique compared to existing pathological speech databases, which do not include phonetic-level annotations. Demographic details such as age, sex, and clinical voice quality (GRBAS score [45]) were also recorded. Markers agreed to omit diacritics except in cases of severe speech impairment, where velarized/dark ‘ë’ was used when necessary. Voiceless, labialized, and nasalized diacritics were also applied in such cases. Additionally, the ‘e’ vowel was transcribed for words such as /bed/, rather than using the ‘Ç’ vowel. Regional accent classifications were noted during the transcription by the transcriber. This was determined through a combined approach of speaker nationality data and acoustic-phonetic analyses performed by trained speech therapists.

##### Dysfluencies

Dysfluencies are widely known to be common in stroke survivors. For consistency, transcribers followed the CHAT conventions when denoting pauses. Pauses shorter than 2.0 seconds were coded as “(.)”. For pauses between 2.01 and 5.00 seconds, the code “(..)” was used. Pauses lasting between 5.01 and 10.00 seconds were transcribed as “(…)”. When the pause exceeded 10.00 seconds, the exact duration was specified, e.g., “(12.0)”.

##### Local Events

Non-verbal communication, including gestures, mimes, and emotional states, was annotated. For example, a participant gesturing to a cat or miming a cat catching a fish was documented. Emotional states during spontaneous speech, such as frustration, anger, and laughter, were also captured in annotations.

##### Error Coding

Transcribers marked four types of language errors: (a) semantic [* s], (b) phonological [* p], (c) morphological [* m], and (d) neologism [* n]. A language error category was further specified by detailing the code if the intention of the speaker was clear e.g., [* s:r] for a semantic error where a related word was used; [* p:w] for a phonological error where a real-word was used, among other specific language errors [46].

##### Additional Analysis

CLAN commands generated morphological (%mor) and grammatical (%gra) tiers for detailed quantitative linguistic analysis. Profiling commands from CLAN enabled the analysis and quantification of specific language measures from the picture description task, for a total of 11 measures. Measures investigated included the total percentage and count of disfluencies (FLUCALC), total word errors and percentage of word errors (EVAL), total utterance errors (EVAL), percentage of flawed syntax sentences (C-NNLA), words per minute and utterances per minute (Time-dur), number of unique words (types), total words (tokens), and type-token ratio (FREQ TTR). The distribution of such measures are reported in the results section as well as in the appendix (Fig. 3).

**Fig. 1.**
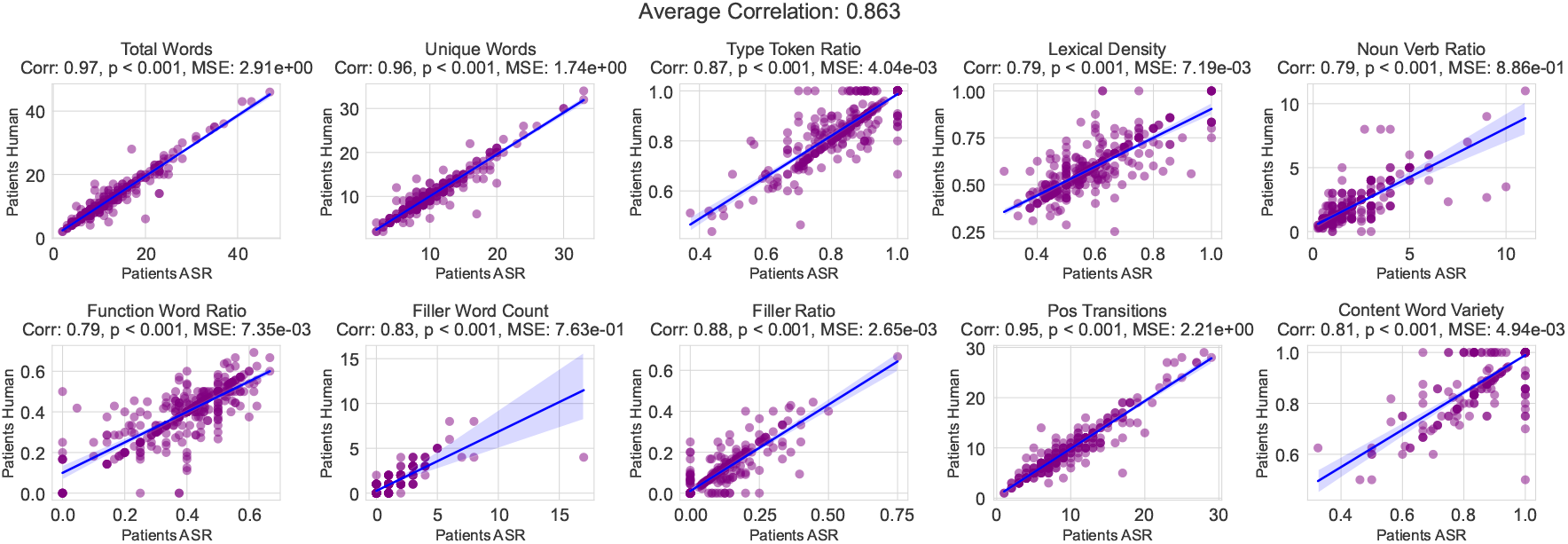
Correlation of linguistic features derived from ASR output and human-labelled speech. For detailed description of the individual linguistic features, see methods. Each point represents a patient (N=576).

**Fig. 2.**
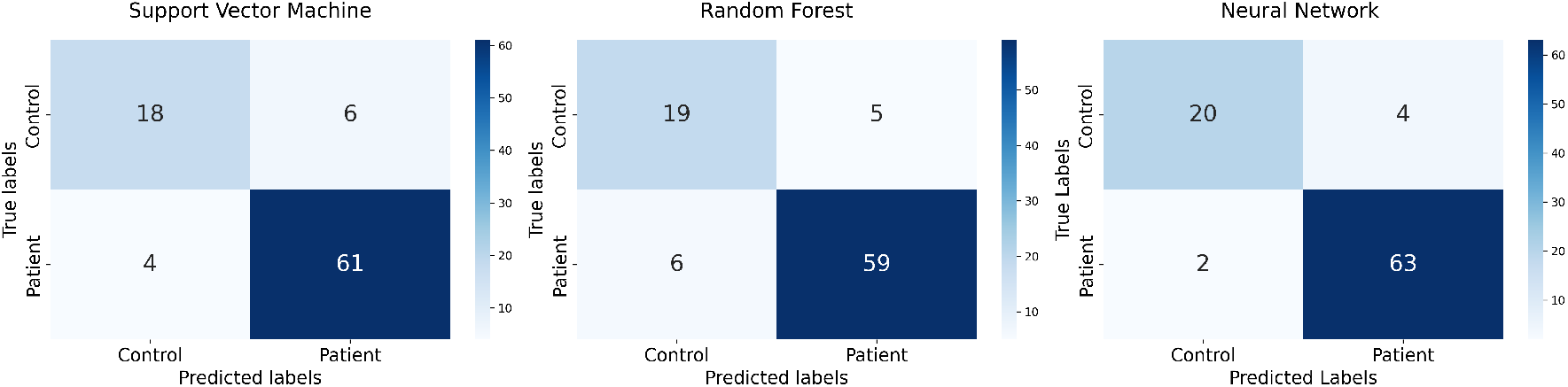
Confusion matrix of the three classification methods used to classify stroke survivors vs. non-stroke healthy controls (N of files in the test set = 65 and 24, respectively), using acoustic features derived from openSMILE. Colourbar represents absolute numbers

**Fig. 3.**
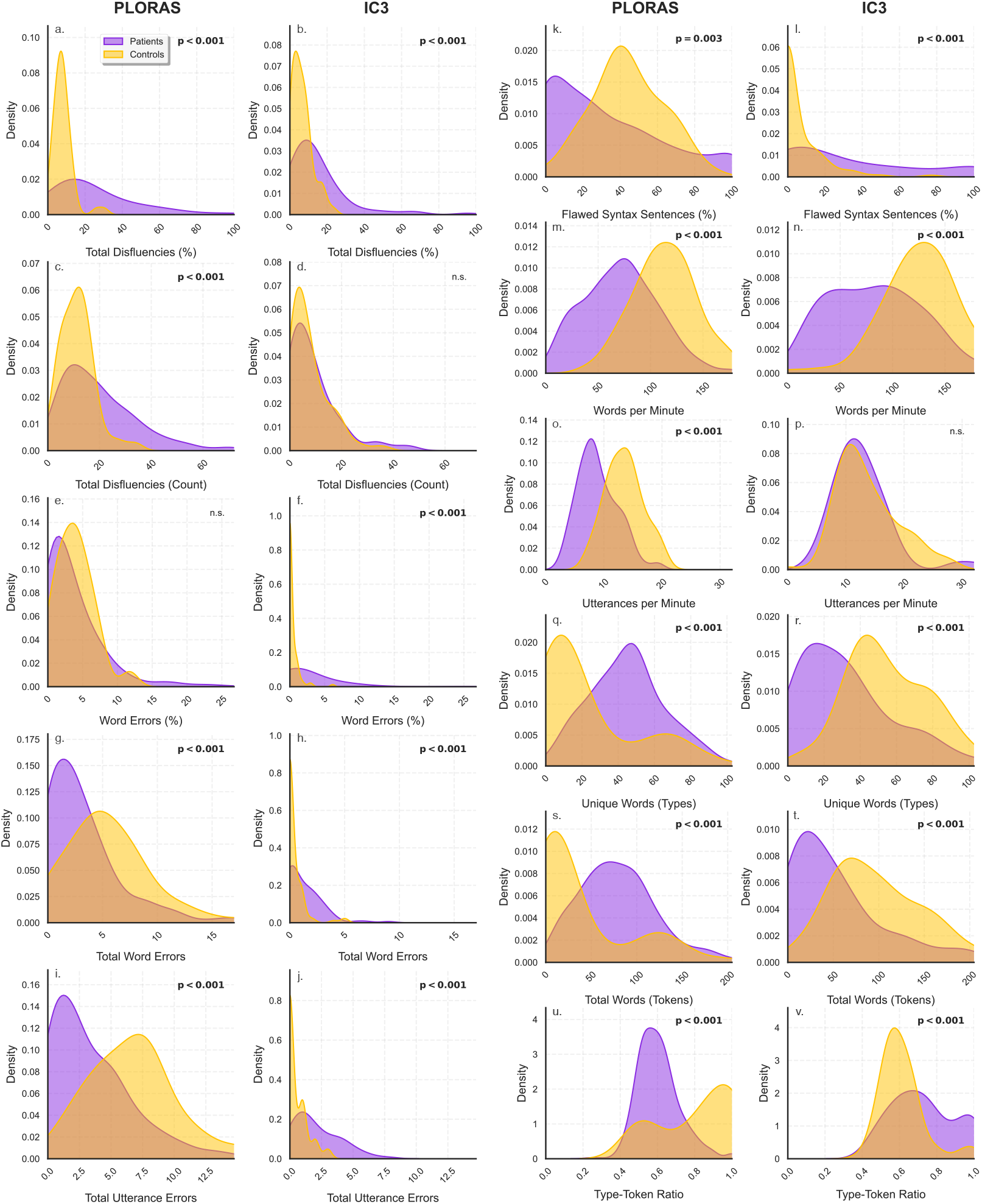
Density plots of CLAN-extracted linguistic features from patient and control transcriptions in the SONIVA database. Data are shown separately for the IC3 (Patients ***N* = 47**, Controls ***N* = 53**) and PLORAS (Patients ***N* = 576**, Controls ***N* = 51**) picture description protocols. Purple and yellow represent patients and controls, respectively. Subplots are labeled (a)–(v), with each pair of vertically arranged plots representing a single linguistic feature across PLORAS (left) and IC3 (right). P-values ***<*** 0.001 are Bonferroni corrected; “n.s.” indicates non-significant differences.

Transcribers initially underwent group training using a shared set of 10 patient files. Afterwards, seven individual transcribers independently annotated a different set of 10 patient files. A two-way mixed-effects intra-class correlation (ICC) analysis was conducted on 15 linguistic measures to assess inter-rater reliability. These measures included both error coding variables (4 features) and the additional linguistic analysis metrics (11 features). The ICC analysis aimed to evaluate consistency across transcribers, ensuring that the independently generated annotations were reliable and comparable. The intra-class correlation analysis revealed a high level of inter-rater reliability for the majority of linguistic measures, with most of those achieving the ‘excellent’ criterion (ICC *>* 0.90). However, five measures did not reach this highest category. Three measures were rated as ‘good’ (ICC between 0.75 and 0.89): *neologism frequency* (FREQ neologism; ICC = 0.860), *total word errors* (EVAL word error total; ICC = 0.767), and *word error percentage* (EVAL word error percentage; ICC = 0.724). One measure, *morphological errors* (FREQ morphology; ICC = 0.661), was rated as ‘moderate/acceptable’ (ICC between 0.50 and 0.74). Lastly, *phonological errors* (FREQ phonology; ICC = 0.311) was rated as ‘poor’ (ICC *<* 0.5).

#### Single-Word Task Annotation

For IC3, single-word tasks (naming, reading, and repetition) were annotated at the trial level. Each trial was scored on a 0, 1, or 2 scale for each measure - semantic (naming task only), dysfluency, self-correction, and phonology. Higher scores indicated better performance. For patient data, speech was transcribed verbatim as ground truth, including fillers such as “er” and non-informative carrying phrases such as “I don’t know, maybe it’s a… “. The timestamp for all participants’ responses was recorded from the onset time of each target word using Audacity editing software (v. 2.7.3) [47]. Technical errors, background noises, and participant skips due to speech impediments were categorised via specifically agreed-upon codes created by trained transcribers.

### C. Fine-tuning ASR on SONIVA

We conducted two experiments to evaluate SONIVA. The first focused on fine-tuning Whisper to extract linguistic features from pathological speech and compare them with goldstandard human transcriptions. The second, detailed in the next section, assessed machine learning classifiers trained on acoustic features to distinguish stroke from control speech.

We selected Whisper [24], a foundation ASR model previously applied in our work [21], [22], [48], due to its robustness across diverse speech conditions and adaptability to low-resource settings. For this experiment, we used the medium-sized Whisper model and fine-tuned it on the annotated portion of SONIVA, consisting of 576 stroke survivors performing the picture description task. The dataset was partitioned into training (70%), validation (18%), and unseen test (12%) sets, stratified by stroke severity and dataset source (IC3 or PLO-RAS) to ensure balanced representation.

Fine-tuning followed our prior methodology [21], [22]. All parameters were updated without layer freezing, using a per-device batch size of 16, gradient accumulation, and a cosine learning rate schedule (starting at 1 *×* 10^−5^ with 1,000-step warm-up). Optimisation used AdamW with cross-entropy loss, evaluated by Word Error Rate (WER) every 1,000 steps. Training was capped at 6,000 steps, with the best model selected based on WER. Mixed-precision (fp16) and gradient checkpointing were applied to improve efficiency. Training completed in 8 hours using PyTorch and Hugging Face Transformers on an NVIDIA RTX 6000 GPU.

Following training, the unseen test set was used exclusively for linguistic feature extraction, allowing for an independent assessment of the model’s ability to generalise. Key linguistic features were extracted and correlated with those extracted from human transcriptions. This step was crucial to assess the reliability of the ASR-based analysis, ensuring that the automatically derived features align closely with those obtained from human transcriptions. Using the NLTK library, commonly used for tokenisation, part-of-speech tagging, and stop word identification [49], we extracted 10 linguistic features categorised into Lexical Diversity, Syntax, and Dysfluency metrics, capturing key aspects of language complexity and fluency.

1. Lexical Diversity metrics include *Total Words* (word count, *N*_words_) and *Unique Words*, (distinct tokens, *N*_unique_), from which we also compute the *Type-Token Ratio* 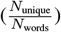, assessing vocabulary variation; and *Lexical Density* 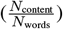 quantifies the proportion of content words (*N*_content_).
2. Syntax structure is evaluated through *Noun-to-Verb Ratio* 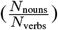; The *Function Word Ratio* 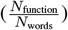, where function words include prepositions, conjunctions, determiners, pronouns, and auxiliary verbs, indicating syntactic cohesion; and *Part-of-speech Transitions* capturing sentence complexity; *Unique Content Word Variety* 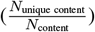 measuring the diversity of meaningful words (nouns, verbs, adjectives, and adverbs) capturing the richness of conveyed information.
3. Dysfluency metrics include *Filler Word Count*, identifying hesitation markers, and *Filler Ratio* 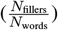, measuring speech fluidity.

Finally, the linguistic features derived automatically and manually were correlated using a Spearman correlation.

### D. Acoustic Feature Analysis

The second experiment aimed to evaluate whether acoustic features extracted from the SONIVA database could reliably distinguish stroke survivors from non-stroke healthy controls. Acoustic features were extracted using the openSMILE Python library (v. 3.0.1) with the extended Geneva Minimalistic Acoustic Parameter Set (eGeMAPSv02) [50]. This standardised feature set comprises 88 parameters, selected for their relevance in clinical and paralinguistic speech analysis, already widely clinically proven for assessing pathological speech in patients with motor speech sound disorders (rather than the aphasic patient population) [51]–[55]. The acoustic feature set encompasses prosodic measures (e.g., *F*_0_ stability, jitter, shimmer) to assess pitch and phonatory control, spectral features (e.g., MFCCs, flux) for vocal resonance and articulatory precision, and energy-related metrics (e.g., loudness, harmonics-to-noise ratio, pauses) to evaluate fluency and rhythmic disturbances. All features were included in the classification models.

Three distinct classification models were employed: (1) Support Vector Machine (SVM) utilising a radial basis function (RBF) kernel, with hyperparameters tuned using grid search to optimise performance, and probability estimates enabled for more informative predictions; (2) Random Forest (RF) comprising 100 trees, with the `sqrt` method for feature selection at each tree bifurcation, and unrestricted tree depth to adapt to the complexity of the data; and (3) Neural Network (NN) using a fully connected architecture designed specifically for this task. The Neural Network consisted of an input layer matching the feature dimensions, followed by two hidden layers of 128 and 64 units, respectively. Each hidden layer employed Leaky ReLU activation, batch normalisation, and dropout (40%) to improve generalisation. The output layer was a single neuron with a sigmoid activation function for binary classification.

Audio files were taken from the SONIVA database (N=576 SS; 104 controls; Total = 680). The corpus was split into a training set (91.03%) and a test set (8.97%) based on total audio duration (see Tab. II). While a stratified approach was used to retain equal proportions of each target class, the final proportion deviated slightly from 90/10 due to variability in audio length across recordings rather than simply the number of files. The training data was further split using a 5-fold group cross-validation strategy to train and validate the models. Due to the longitudinal aspect of the SONIVA data collection, where some patients had multiple recordings over time, the cross-validation was structured in groups (`StratifiedGroupKFold`) to restrict each unique patient data within either the training or validation folds, avoiding information leakage. This approach also guaranteed that the external test set comprised only previously unseen patients. To address class imbalance, data augmentation was performed on the minority class (non-stroke control) using SMOTE [56] in the training set, while the test set remained unmodified to ensure a realistic evaluation. Feature standardisation was performed within each fold, using only training data to prevent data leakage. Evaluation metrics such as accuracy and F1-score were tested on the test set before created.

## II. RESULTS

### A. SONIVA data collection

Speech data from ≈1000 patients with stroke, including *>* 200 with longitudinal data, have been collected. The manually transcribed component of the larger SONIVA database has 26,783 files to date, comprising recordings from 618 stroke survivors and 189 non-stroke control group (Table III). Picture description tasks contribute to 825 transcriptions, while single-word tasks account for 25,429 transcriptions, providing a diverse linguistic and articulatory representation. Speech recordings from an additional 155 stroke survivors, available but not yet fully transcribed, are not listed in this table. Figure 3 shows the distribution of linguistic features extracted from CLAN for those manually transcribed to date.

**TABLE III.** Overview of the annotated SONIVA database composition.

| Task Type | Dataset | Controls (files) | Patients (files) | Total Transcriptions |
| --- | --- | --- | --- | --- |
| Picture Description | IC3<sup>†</sup> | 53<br>(CAT=65; Beach=66) | 47<br>(CAT=57; Beach=57) | 245 |
|  | PLORAS | 51 | 529 | 580 |
|  | **Total** | **104** | **576** | **825** |
| Naming | IC3 | 138 (8,371) | 89 (4,200) | 12,571 |
| Reading | IC3 | 138 (3,038) | 88 (1,518) | 4,556 |
| Repetition | IC3 | 138 (5,582) | 88 (2,720) | 8,302 |
|  | **Total** | **138** | **89** | **25,429** |
|  | **GRAND TOTAL<sup>‡</sup>** | **189<sup>†</sup>** | **618<sup>†</sup>** | **26,254** |

<sup>†</sup>All IC3 picture description participants also completed the single word tasks
<sup>‡</sup>Grand total accounts for overlap between IC3 picture description and single word participants

The age and gender demographics of stroke survivors in SONIVA align with typical stroke population patterns that commonly take part in research trials (Fig. 4). Male speakers constitute the majority (69.81%) of the database, averaging 62.91 years, while female speakers average 57.35 years at testing. This gender imbalance reflects global stroke incidence patterns, particularly in younger age groups where males have higher stroke occurrence rates [57].

**Fig. 4.**
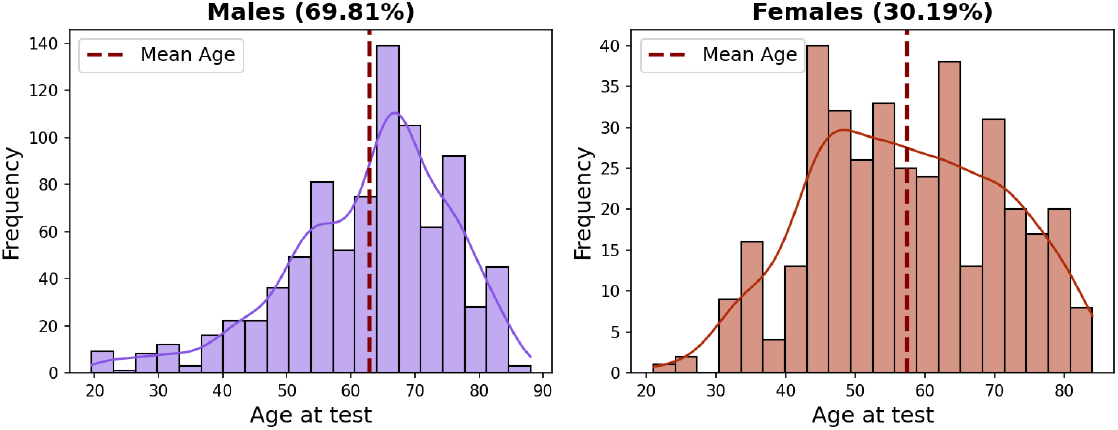
Distribution of age and gender across stroke survivors in the SONIVA database as shown in Tab. 3.

SONIVA includes tasks conducted in British English. However, given the clinical nature of the database derived from a metropolitan population, it naturally captures a diverse range of accents, reflecting its broad geographical representation (Fig. 6). This is important to capture, as accents and regional dialects can significantly affect the generalisability of language and acoustic models [58]. The majority of speakers (76.77%) have UK-based regional accents, with the remainder spanning multiple regions: European (2.99%), South Asian (3.67%), African (2.85%), Central American (2.58%), Oceanian (0.68%), and North American (0.27%). Among UK-based speakers (Fig. 5), 53.2% exhibit “Standard British accents”, a designation encompassing neutral English accents and speakers without a strong regional affiliation. 22.8% of participants have specific London-area accents, but the remainder includes South East England (5.8%), Scotland (5.7%), Yorkshire, and The Humber (5.5%), East Midlands (3.6%), and West Midlands (3.4%). Less frequently represented accents include South West England (2.9%), Northern Ireland (2.7%), Wales (1.6%), North West England (1.3%), East of England (0.4%), and North East England (0.4%), bringing a broad phonetic and linguistic diversity into the dataset. A further 10.19% of speakers presented mixed or unclassified accents that did not fit within the primary categories.

**Fig. 5.**
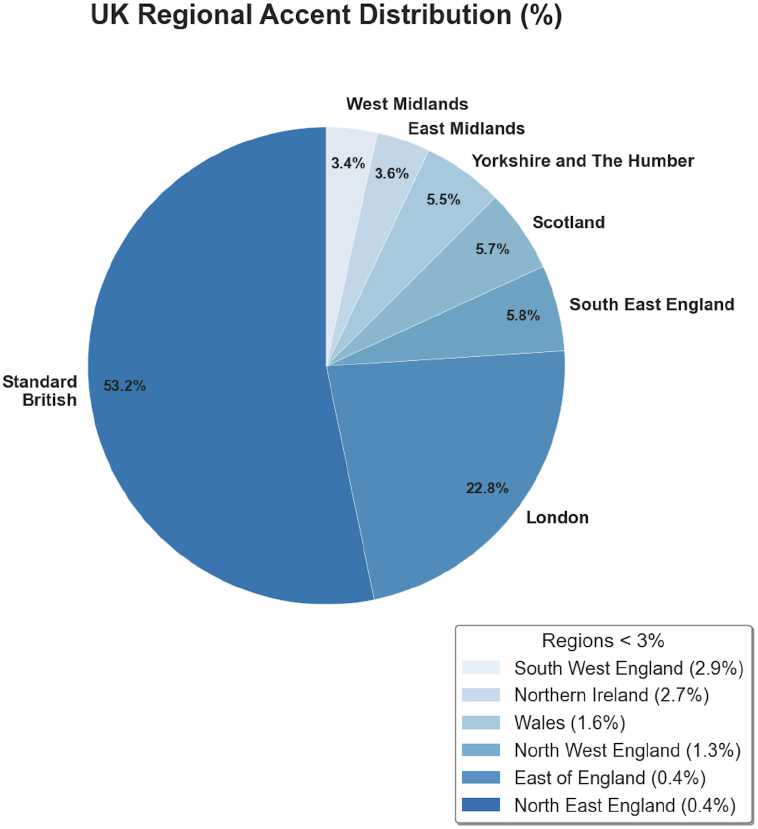
UK accent distribution across stroke survivors in the SONIVA database as shown in Tab. 3.

**Fig. 6.**
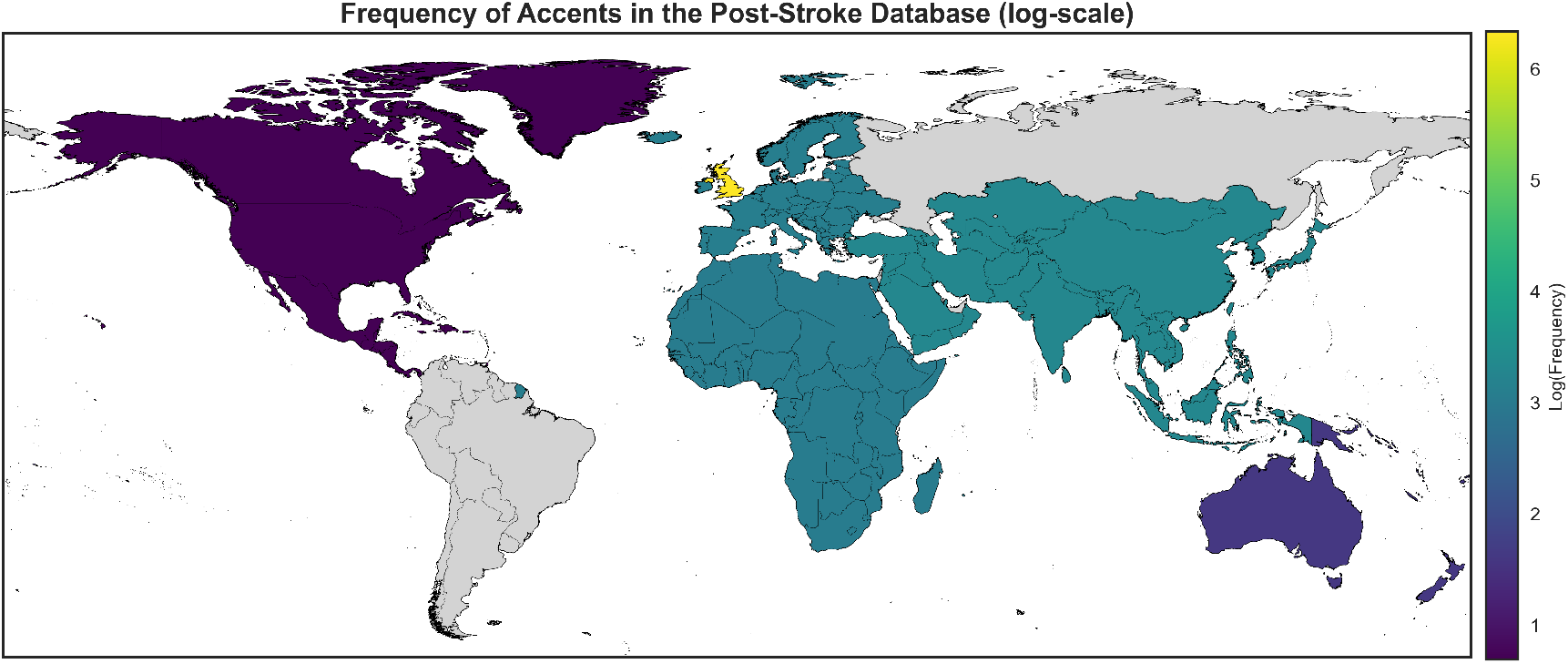
World accent distribution across stroke survivors in the SONIVA database as shown in Tab. 3.

### B. Experiments Result

Fine-tuning the ASR significantly improved its performance, reducing WER from 39.60% to 21.51% on the validation set and from 43.62% to 21.93% on the test set. Further analysis comparing ASR-derived linguistic features with those derived from human transcriptions revealed robust correlations across all measured variables (see results in Fig. 1), yielding an average correlation of 0.863. Basic lexical measures demonstrated strong correlations, with total words, unique words, and type-token ratio showing correlations of 0.97, 0.96, and 0.87, respectively, accompanied by low mean squared errors (MSE: 2.91e+00, 1.74e+00, and 4.04e-03). Lexical density also exhibited notable alignment (Corr: 0.79, MSE: 7.19e-03). Grammatical structure metrics showed high reliability, with function word ratio (Corr: 0.79, MSE: 7.35e-03), part-of-speech transitions (Corr: 0.95, MSE: 2.21e+00), and noun-verb ratio (Corr: 0.79, MSE: 8.86e-01). Measures of disfluency and content analysis similarly displayed strong alignment, with filler word count (Corr: 0.83, MSE: 7.63e-01), filler ratio (Corr: 0.88, MSE: 2.65e-03), and content word variety (Corr: 0.81, MSE: 4.94e-03). Overall, these results support the feasibility of using ASR-based tools for automated linguistic analysis in clinical applications using the SONIVA database. Figure 2 and Table IV present the classification performance of Support Vector Machine (SVM), Random Forest (RF), and Neural Network (NN) models on the SONIVA dataset using acoustic features. The confusion matrices illustrate the models’ ability to distinguish stroke survivors from non-stroke controls, while Table IV provides precision, recall, and F1-scores. Among the models, NN achieved the highest classification accuracy of 93%, followed by SVM and RF with 89% and 88%, respectively. The NN model performed best, achieving 0.92 precision and 0.90 recall, resulting in an F1-score of 0.91. SVM and RF exhibited similar F1-scores of 0.85, with SVM achieving a slightly higher precision of 0.86 compared to RF’s 0.84. The confusion matrices in Fig. 2 confirm that NN produced the fewest misclassifications, while RF and SVM showed slightly lower recall, misclassifying a marginally higher number of stroke participants. Overall, the results indicate that the NN model provided the most robust classification performance, achieving the highest accuracy and F1-score, making it the most effective classifier for distinguishing between stroke survivors and healthy controls.

**TABLE IV.**
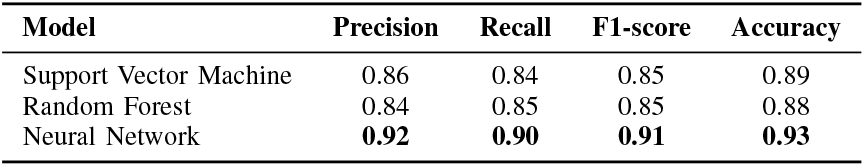
Performances of the three classification methods using openSMILE acoustic features.

## III. DISCUSSION

This study introduces the SONIVA database and validates its utility for analysing stroke-induced speech impairments. The findings from both linguistic and acoustic feature analyses provide strong evidence of its value in capturing meaningful speech characteristics for clinical applications, such as disease classification and automated speech analysis. SONIVA represents the largest stroke survivors’ speech database to date, and uniquely, it includes approximately 200 patients with longitudinal assessments. This extensive follow-up is crucial for tracking recovery trajectories, understanding individual variability in post-stroke outcomes, and identifying predictive markers of long-term cognitive and language impairments. To the best of the authors’ knowledge, SONIVA is the most comprehensive dataset of its kind, offering an unprecedented opportunity to study both cross-sectional and longitudinal aspects of speech and cognitive recovery following stroke.

Whisper fine-tuning significantly improved the WER performance, reducing it ≈20% (from 43.62% to 21.93% on the test set), demonstrating the benefits of domain-specific adaptation of existing attention-based ASR models using our pathological speech database. Prior work using MT-DNN with BLSTM [29], achieved a WER of 42.4% for impaired speech and of 18.5% for healthy speech [26], [59], while more recent E-Branchformer and Conformer models [30] reported an average WER of 26% across various aphasia severity levels. In contrast, our approach demonstrates better performance, achieving significantly lower WER for pathological speech, reinforcing the value of SONIVA in fine-tuning strategies in clinical ASR applications. We have also previously shown the generalisation capability of such models across diverse datasets [21], beyond SONIVA. We demonstrated that finetuned ASR on the SONIVA database can improve ASR performance applied to other databases such as the AphasiaBank (with different language tasks and accents) and also different pathologies, such as dementia (DementiaBank).

Beyond WER improvements, fine-tuned ASR-derived linguistic features strongly correlated with human-transcribed features, confirming SONIVA’s potential to enhance automated speech analysis in stroke populations. These correlations suggest that ASR-based tools can provide clinically relevant linguistic metrics, reducing reliance on manual transcription for assessment and monitoring.

The classification models based on acoustic features further highlight the value of large-scale speech datasets in developing automated diagnostic tools. The NN model achieved the highest accuracy (93%), with a precision of 0.92 and recall of 0.90, outperforming SVM (89% accuracy) and RF (88%). While SVM showed slightly higher precision (0.86) than RF (0.84), their recall values were similar. The NN’s superior performance suggests that deep learning approaches, when optimised, can surpass traditional machine learning methods in pathological speech classification.

These findings align with or surpass previous studies focusing on speech classification in patients with dysarthria or other laryngeal pathologies, where the classification method has heavily relied on acoustic features predominantly extracted through openSMILE. For instance, [52] analysed the vowels /a/, /i/, and /u/ under normal, high, low, and rising-falling pitch conditions, reporting classification accuracies of 0.82 with the ComParE feature set and 0.76 with the eGeMAPS set when distinguishing between various voice disorders; spasmodic dysphonia, recurrent laryngeal nerve palsy, functional dysphonia, and psychogenic dysphonia. When detecting dysarthria in speech data from Torgo and UA-Speech in [60] work and Saarbrücken database in [61] work, accuracies of 0.92 have been reported for distinguishing healthy speakers and individuals with voice pathologies such as pachydermia, laryngitis, spasmodic dysphonia or vocal fold paresis.

However, these databases primarily contain short speech samples or isolated vowels, limiting their applicability for language-based assessments. Our prior work [22] showed that language models perform poorly on such datasets due to their lack of continuous speech. It is not surprising that our significantly larger dataset (*N >* 400 compared to *<* 15 in TORGO and UA-Speech) provides greater statistical power and better representation of language variations, allowing for more robust language model training and generalisation for language-based aphasia classification [22]. It can therefore be concluded that the development of large, long-form speech databases is essential for enabling the classification of more fine-grained aphasia subtypes, providing a deeper and more precise understanding of linguistic impairments [31].

Whilst SONIVA shows promising results for clinical application and pathological speech research, certain challenges remain. The presence of background clinical environmental noise and overlapping speech in the recordings introduce additional complexity in model learning [62]. Future improvements could incorporate speaker diarisation, segmentation and voice activity detection techniques. Moreover, performance of the ASR and acoustic models could have been improved further by exploring additional hyper-parameter fine-tuning or adopting alternative architectures altogether.

Although the SONIVA database encompasses stroke survivors with a wide range of stroke and aphasia severity, this initial effort in developing ASR prioritised human transcription for the most severely impaired individuals. The data from the IC3 and PLORAS database had different clinical ratings for severity assessment. In the IC3 dataset, the NIHSS scale [41] was used as the primary measure of overall stroke severity (mean = 3.29; range = 0–13). For the PLORAS database, clinical aphasia severity was determined by speech and language pathologists using the picture-description subtest from the Comprehensive Aphasia Test (CAT), as previously defined (mean = 28.20; range = 0–85). In future, a unified clinical severity score across individuals in the two different arms of the SONIVA database will allow for a better evaluation of any developed classification models.

Beyond advancing models for pathological disease classification and severity assessment, SONIVA offers a powerful framework for detecting specific aphasic linguistic impairments, such as semantic and phonological errors. This remains a critical yet unresolved challenge in the field, positioning SONIVA as a novel tool for addressing a longstanding gap in aphasia detection and analysis. To this end, the International Phonetic Alphabet (IPA) transcriptions present in SONIVA enable detailed phonetic error analysis, paving the way for a phonetic diagnostic platform. Additionally, paralinguistic feature annotations and linked self-rated mood questionnaires can support predictive models for emotional well-being, frequently affected post-stroke [63], [64].

Since patients enrolled in the SONIVA dataset have linked neurobiological data, such as clinical information, general cognitive scores, and MRI brain scans, this provides the opportunity to use a priori information about brain health to make predictions about linguistic outcomes after injury [4]. Equally, speech derived metrics can speech-derived metrics can also serve as predictors of general cognitive function after brain injury [65], reinforcing the bidirectional relationship between language processing and neurological status.

Finally, SONIVA’s comprehensive dataset advances both research and clinical applications that goes beyond post-stroke aphasia, highlighting its translational potential. Indeed, its utility extends to studying speech disorders in different types of dementia, motor neuron disease or traumatic brain injuries. This is because, as mentioned above (see Sec.) stroke-related speech impairments exhibit significant heterogeneity, encompassing a broad spectrum of motor and linguistic deficits. These characteristics mirror the diverse speech disturbances observed in other neurological conditions, making SONIVA a valuable resource for developing models that generalise across different disorders. This cross-pathology generalisation, already proved in other preliminary results still using SONIVA and foundation models [22], enables comparative analyses that may uncover shared mechanisms and disorder-specific features, making SONIVA a valuable resource for broader neurological speech research.

## IV. CONCLUSION

The findings underscore SONIVA’s value as a foundational resource for post-stroke speech pathology research for future advancements and refinements. Its combination of linguistic and acoustic analyses, strong classification performance, and longitudinal design makes it well-suited for advancing both ASR-based and acoustic feature-based assessments. Furthermore, its framework has promising implication that extends beyond its immediate application in stroke-related speech analysis, potentially able to offer a template for broader applications in speech pathology research different from stroke. This work lays the foundation for scalable, automated speech pathology solutions, aiming to integrate SONIVA into digital health technologies, remote monitoring, and personalised therapy to enhance patient outcomes and support AI-driven clinical decision systems.

## AUTHOR CONTRIBUTION

G.S. developed the computational pipeline, performed data preprocessing and analysis (including ASR fine-tuning, NLP feature extraction, and classification tasks), and wrote the manuscript. F.G. and P.A.N. supervised the study and provided guidance on experimental design, clinical interpretation, and manuscript development. C.J.P. and F.G. were responsible for the oversight of the PLORAS and IC3 studies respectively. S.B., D.C.G., and N.V.P. managed the collection, marking and quality control of the IC3 dataset. All authors contributed to the manuscript. All authors declare no financial or non-financial competing interests.

## DATA AVAILABILITY

The speech-derived linguistic features of SONIVA are available at: https://github.com/Clinical-Language-Cognition-Lab/SONIVA_paper^3^. The full SONIVA database is available to academic researchers upon request, subject to standard institutional regulatory approvals and data sharing agreements.

## CODE AVAILABILITY

The SONIVA fine-tuned Whisper ASR model is available at: https://github.com/Clinical-Language-Cognition-Lab/SONIVA_paper^4^.

## INCLUSION & ETHICS STATEMENT

All data collection protocols received ethical approval from the UK Health Research Authority (IRAS: IC3-299333 and PLORAS-133939), and informed consent was obtained from all participants. The SONIVA database reflects a diverse clinical population, including a broad range of age, gender, regional accents, and neurological severity, supporting inclusive research and model generalizability. No personally identifiable information is shared, and all data handling complies with institutional and GDPR standards.

## ACKNOWLEDGMENTS

The authors would like to thank A. Coghlan, O. Burton and N. Parkinson for assistance with the IC3 study, and J. Friedland, K. Stephenson, G. Gvero, Z. Zaskorska and C. Leong for labelling the speech data. We are grateful to the patients, clinical teams, and the PLORAS team for their valuable contributions to the SONIVA database. Infrastructure support was provided by the NIHR Imperial Biomedical Research Centre and the NIHR Imperial Clinical Research Facility. The views expressed are those of the authors and not necessarily those of the NHS, the NIHR or the Department of Health and Social Care. Funding support was provided to G.S. (UK Research and Innovation [UKRI Centre for Doctoral Training in AI for Healthcare:EP/S023283/1]), F.G. (Medical Research Council P79100) and S.B. (EPSRC IAA -PSP415 and MRC IAA -PSP518).

APPENDIX

^1^IC3: IRAS 299333, registered on ClinicalTrials.gov (NCT05885295); PLORAS: IRAS 133939, REC reference 13/LO/1515.

^2^https://www.cognitron.co.uk.

^3^The repository will be populated prior to final review or upon request during earlier stages of peer review.

^4^The repository will be populated prior to final review or upon request during earlier stages of peer review.

## Notes

### Competing Interest Statement

The authors have declared no competing interest.

### Clinical Protocols

https://clinicaltrials.gov/study/NCT05885295

### Author Declarations

Both IC3 and PLORAS studies received ethical approval from the UK Health Research Authority - IC3: IRAS 299333, registered on ClinicalTrials.gov (NCT05885295) - PLORAS: IRAS 133939, REC reference 13/LO/1515

## REFERENCES

[1] H. L. Flowers, F. L. Silver, J. Fang, E. Rochon, and R. Martino, “The incidence, co-occurrence, and predictors of dysphagia, dysarthria, and aphasia after first-ever acute ischemic stroke.” Journal of communication disorders, vol. 46, no. 3, pp. 238–248, 2013.

[2] C. Ellis, A. N. Simpson, H. Bonilha, P. D. Mauldin, and K. N. Simpson, “The one-year attributable cost of poststroke aphasia.” Stroke, vol. 43, no. 5, pp. 1429–1431, 2012.

[3] G. Hill, S. Regan, R. Francis, G. Mead, S. Thomas, R. A.-S. Salman, C. Roffe, A. Pollock, S. Davenport, E. Kontou et al., “Research priorities to improve stroke outcomes.” The Lancet Neurology, vol. 21, no. 4, pp. 312–313, 2022.

[4] J. D. Stefaniak, F. Geranmayeh, and M. A. Lambon Ralph, “The multidimensional nature of aphasia recovery post-stroke.” Brain, vol. 145, no. 4, pp. 1354–1367, 2022.

[5] J. S. Damico, N. Müller, and M. J. Ball, The handbook of language and speech disorders. Wiley Online Library, 2010.

[6] J. Fridriksson, H. I. Hubbard, S. G. Hudspeth, A. L. Holland, L. Bonilha, D. Fromm, and C. Rorden, “Speech entrainment enables patients with broca’s aphasia to produce fluent speech.” Brain, vol. 135, no. 12, pp. 3815–3829, 2012.

[7] R. A. Butler, M. A. Lambon Ralph, and A. M. Woollams, “Capturing multidimensionality in stroke aphasia: mapping principal behavioural components to neural structures.” Brain, vol. 137, no. 12, pp. 3248–3266, 2014.

[8] G. Fergadiotis, K. Gorman, and S. Bedrick, “Algorithmic classification of five characteristic types of paraphasias.” American Journal of Speech-Language Pathology, vol. 25, no. 4S, pp. S776–S787, 2016.

[9] E. Charters, S. Coulson, and T. Low, “Oral incompetence: Changes in speech intelligibility following facial nerve paralysis.” J. of Plastic, Reconstructive & Aesthetic Surgery, vol. 87, pp. 472–478, 2023.

[10] K. Knollman-Porter, “Acquired apraxia of speech: a review.” Topics in stroke rehabilitation, vol. 15, no. 5, pp. 484–493, 2008.

[11] R. Lorenz, M. Johal, F. Dick, A. Hampshire, R. Leech, and F. Geranmayeh, “A bayesian optimization approach for rapidly mapping residual network function in stroke.” Brain, vol. 144, no. 7, pp. 2120–2134, 2021.

[12] F. Geranmayeh, R. Leech, and R. J. Wise, “Network dysfunction predicts speech production after left hemisphere stroke.” Neurology, vol. 86, no. 14, pp. 1296–1305, 2016.

[13] F. Geranmayeh, T. W. Chau, R. J. Wise, R. Leech, and A. Hampshire, “Domain-general subregions of the medial prefrontal cortex contribute to recovery of language after stroke.” Brain, vol. 140, no. 7, pp. 1947–1958, 2017.

[14] F. Geranmayeh, R. J. Wise, A. Mehta, and R. Leech, “Overlapping networks engaged during spoken language production and its cognitive control.” Journal of Neuroscience, vol. 34, no. 26, pp. 8728–8740, 2014.

[15] F. Geranmayeh, S. L. Brownsett, and R. J. Wise, “Task-induced brain activity in aphasic stroke patients: what is driving recovery?” Brain, vol. 137, no. 10, pp. 2632–2648, 2014.

[16] S. Spaccavento, A. Craca, M. Del Prete, R. Falcone, A. Colucci, A. Di Palma, and A. Loverre, “Quality of life measurement and outcome in aphasia.” Neuropsychiatric disease and treatment, vol. 10, p. 27, 2014.

[17] S. Roberts, R. M. Bruce, L. Lim, H. Woodgate, K. Ledingham, S. Anderson, D. L. Lorca-Puls, A. Gajardo-Vidal, A. P. Leff, T. M. Hope et al., “Better long-term speech outcomes in stroke survivors who received early clinical speech and language therapy: What’s driving recovery?” Neuropsychological Rehabilitation, vol. 32, no. 9, pp. 2319–2341, 2022.

[18] M. C. Brady, H. Kelly, J. Godwin, P. Enderby, and P. Campbell, “Speech and language therapy for aphasia following stroke.” Cochrane database of systematic reviews, no. 6, 2016.

[19] D. Le, K. Licata, and E. M. Provost, “Automatic quantitative analysis of spontaneous aphasic speech.” Speech Communication, vol. 100, pp. 1–12, 2018.

[20] R. Palmer, P. Enderby, C. Cooper, N. Latimer, S. Julious, G. Paterson, M. Dimairo, S. Dixon, J. Mortley, R. Hilton et al., “Computer therapy compared with usual care for people with long-standing aphasia post-stroke: a pilot randomized controlled trial.” Stroke, vol. 43, no. 7, pp. 1904–1911, 2012.

[21] G. Sanguedolce, S. Brook, D. C. Gruia, P. A. Naylor, and F. Geranmayeh, “When whisper listens to aphasia: Advancing robust post-stroke speech recognition.” in Proc. of Interspeech, 2024, pp. 1995–1999.

[22] G. Sanguedolce, D.-C. Gruia, S. Brook, P. Naylor, and F. Geranmayeh, “Universal speech disorder recognition: towards a foundation model for cross-pathology generalisation.” in Advancements In Medical Foundation Models: Explainability, Robustness, Security, and Beyond, 2024.

[23] S. Hu, S. Liu, X. Xie, M. Geng, T. Wang, S. Hu, M. Cui, X. Liu, and H. Meng, “Exploiting cross domain acoustic-to-articulatory inverted features for disordered speech recognition.” in ICASSP 2022-2022 IEEE International Conference on Acoustics, Speech and Signal Processing (ICASSP). IEEE, 2022, pp. 6747–6751.

[24] A. Radford, J. W. Kim, T. Xu, G. Brockman, C. McLeavey, and I. Sutskever, “Robust speech recognition via large-scale weak supervision.” pp. 28492–28518, 2023.

[25] S. S. Mahmoud, R. F. Pallaud, A. Kumar, S. Faisal, Y. Wang, and Q. Fang, “A comparative investigation of automatic speech recognition platforms for aphasia assessment batteries.” Sensors, vol. 23, no. 2, p. 857, 2023.

[26] D. Le and E. M. Provost, “Improving automatic recognition of aphasic speech with aphasiabank.” in Interspeech, 2016, pp. 2681–2685.

[27] K. C. Fraser, F. Rudzicz, N. Graham, and E. Rochon, “Automatic speech recognition in the diagnosis of primary progressive aphasia.” in Proceedings of the fourth workshop on speech and language processing for assistive technologies, 2013, pp. 47–54.

[28] B. MacWhinney, D. Fromm, M. Forbes, and A. Holland, “Aphasiabank: Methods for studying discourse.” Aphasiology, vol. 25, no. 11, pp. 1286–1307, 2011.

[29] Y. Liu, Y. Qin, S. Feng, T. Lee, and P. Ching, “Disordered speech assessment using kullback-leibler divergence features with multi-task acoustic modeling.” in 2018 11th International Symposium on Chinese Spoken Language Processing (ISCSLP). IEEE, 2018, pp. 61–65.

[30] J. Tang, W. Chen, X. Chang, S. Watanabe, and B. MacWhinney, “A new benchmark of aphasia speech recognition and detection based on e-branchformer and multi-task learning.” arXiv preprint arXiv:2305.13331, 2023.

[31] M. Zusag, L. Wagner, and T. Bloder, “Careful Whisper - leveraging advances in automatic speech recognition for robust and interpretable aphasia subtype classification.” in Interspeech 2023, 2023, pp. 3013–3017.

[32] H. Kim, M. H. Johnson, J. Gunderson, A. Perlman, T. Huang, K. Watkin, S. Frame, H. V. Sharma, and X. Zhou, “UAspeech,” 2023. [Online]. Available: 10.21227/f9tc-ab45

[33] F. Rudzicz, A. K. Namasivayam, and T. Wolff, “The TORGO database of acoustic and articulatory speech from speakers with dysarthria.” Language resources and evaluation, vol. 46, pp. 523–541, 2012.

[34] G. Schu, P. Janbakhshi, and I. Kodrasi, “On using the UA-speech and TORGO databases to validate automatic dysarthric speech classification approaches.” in ICASSP 2023-2023 IEEE International Conference on Acoustics, Speech and Signal Processing (ICASSP). IEEE, 2023, pp. 1–5.

[35] C. Lea, V. Mitra, A. Joshi, S. Kajarekar, and J. P. Bigham, “Sep-28k: A dataset for stuttering event detection from podcasts with people who stutter.” in ICASSP 2021-2021 IEEE International Conference on Acoustics, Speech and Signal Processing (ICASSP). IEEE, 2021, pp. 6798–6802.

[36] D. C. Gruia, V. Giunchiglia, A. Coghlan, S. Brook, S. Banerjee, J. Kwan, P. J. Hellyer, A. Hampshire, and F. Geranmayeh, “Online monitoring technology for deep phenotyping of cognitive impairment after stroke.” medRxiv, 2024.

[37] D.-C. Gruia, W. Trender, P. Hellyer, S. Banerjee, J. Kwan, H. Zetterberg, A. Hampshire, and F. Geranmayeh, “IC3 protocol: a longitudinal observational study of cognition after stroke using novel digital health technology.” BMJ open, vol. 13, no. 11, p. e076653, 2023.

[38] M. L. Seghier, E. Patel, S. Prejawa, S. Ramsden, A. Selmer, L. Lim, R. Browne, J. Rae, Z. Haigh, D. Ezekiel et al., “The ploras database: a data repository for predicting language outcome and recovery after stroke..” Neuroimage, vol. 124, pp. 1208–1212, 2016.

[39] Z. S. Nasreddine, N. A. Phillips, V. Bédirian, S. Charbonneau, V. Whitehead, I. Collin, J. L. Cummings, and H. Chertkow, “The montreal cognitive assessment, MoCA: a brief screening tool for mild cognitive impairment.” Journal of the American Geriatrics Society, vol. 53, no. 4, pp. 695–699, 2005.

[40] J. P. Broderick, O. Adeoye, and J. Elm, “Evolution of the modified rankin scale and its use in future stroke trials.” Stroke, vol. 48, no. 7, pp. 2007–2012, 2017.

[41] L. K. Kwah and J. Diong, “National institutes of health stroke scale (NIHSS).” Journal of physiotherapy, 2014.

[42] K. Swinburn, G. Porter, and D. Howard, “Comprehensive aphasia test.” APA PsycTests, 2004.

[43] G. Conti-Ramsden, “CLAN (Computerized Language Analysis).” Child Language Teaching and Therapy, vol. 12, no. 3, pp. 345–349, 1996.

[44] B. MacWhinney, The CHILDES project: Tools for analyzing talk, Volume II: The database. Psychology Press, 2014.

[45] M. Hirano, “Clinical examination of voice.” 1981.

[46] B. MacWhinney, “Tools for analyzing talk part 2: The clan program.” Talkbank. org, no. 2000, 2017.

[47] A. Team, “Audacity.” 2023, free and open-source audio editor and recorder. [Online]. Available: https://www.audacityteam.org/

[48] G. Sanguedolce, D.-C. Gruia, P. Naylor, and F. Geranmayeh, “Latent representation encoding and multimodal biomarkers for post-stroke speech assessment,” in ICLR 2025 Workshop on World Models: Understanding, Modelling and Scaling.

[49] S. Bird, E. Klein, and E. Loper, Natural language processing with Python: analyzing text with the natural language toolkit. “O’Reilly Media, Inc.”, 2009.

[50] F. Eyben, K. R. Scherer, B. W. Schuller, J. Sundberg, E. André, C. Busso, L. Y. Devillers, J. Epps, P. Laukka, S. S. Narayanan et al., “The geneva minimalistic acoustic parameter set (gemaps) for voice research and affective computing.” IEEE transactions on affective computing, vol. 7, no. 2, pp. 190–202, 2015.

[51] M. Shahin, B. Ahmed, D. V. Smith, A. Duenser, and J. Epps, “Automatic screening of children with speech sound disorders using paralinguistic features.” in 2019 ieee 29th international workshop on machine learning for signal processing (mlsp). IEEE, 2019, pp. 1–5.

[52] P. Barche, K. Gurugubelli, and A. K. Vuppala, “Towards automatic assessment of voice disorders: A clinical approach.” in INTERSPEECH, 2020, pp. 2537–2541.

[53] Y. Liu, M. K. Reddy, N. Penttilä, T. Ihalainen, P. Alku, and O. Räsänen, “Automatic assessment of parkinson’s disease using speech representations of phonation and articulation.” IEEE/ACM Transactions on Audio, Speech, and Language Processing, vol. 31, pp. 242–255, 2022.

[54] C. O. Mawalim, B. A. Titalim, S. Okada, and M. Unoki, “Non-intrusive speech intelligibility prediction using an auditory periphery model with hearing loss.” Applied Acoustics, vol. 214, p. 109663, 2023.

[55] D. Kumar, U. Satija, and P. Kumar, “Pathological speech and electroglot-tography signals analysis using invariance scattering network.” Circuits, Systems, and Signal Processing, pp. 1–18, 2024.

[56] N. V. Chawla, K. W. Bowyer, L. O. Hall, and W. P. Kegelmeyer, “Smote: synthetic minority over-sampling technique.” Journal of artificial intelligence research, vol. 16, pp. 321–357, 2002.

[57] P. Appelros, B. Stegmayr, and A. Terént, “Sex differences in stroke epidemiology: a systematic review.” Stroke, vol. 40, no. 4, pp. 1082–1090, 2009.

[58] A. Yadavalli, G. S. Mirishkar, and A. Vuppala, “Exploring the effect of dialect mismatched language models in telugu automatic speech recognition.” in Proceedings of the 2022 Conference of the North American Chapter of the Association for Computational Linguistics: Human Language Technologies: Student Research Workshop, 2022, pp. 292–301.

[59] D. Le, K. Licata, and E. M. Provost, “Automatic paraphasia detection from aphasic speech: A preliminary study.” in Interspeech, 2017, pp. 294–298.

[60] F. Javanmardi, S. R. Kadiri, and P. Alku, “Pre-trained models for detection and severity level classification of dysarthria from speech.” Speech Communication, vol. 158, p. 103047, 2024.

[61] M. Huckvale, Z. Liu, and C. Buciuleac, “Automated voice pathology discrimination from audio recordings benefits from phonetic analysis of continuous speech.” Biomedical Signal Processing and Control, vol. 86, p. 105201, 2023.

[62] S. Kar, P. Mishra, J. Lin, M.-J. Woo, N. Deas, C. Linduff, S. Niu, Y. Yang, J. McClendon, D. H. Smith et al., “Systematic evaluation and enhancement of speech recognition in operational medical environments.” in 2021 International Joint Conference on Neural Networks (IJCNN). IEEE, 2021, pp. 1–8.

[63] M. L. Hackett, C. Yapa, V. Parag, and C. S. Anderson, “Frequency of depression after stroke: a systematic review of observational studies.” Stroke, vol. 36, no. 6, pp. 1330–1340, 2005.

[64] J.-M. Annoni, F. Staub, L. Bruggimann, S. Gramigna, and J. Bogousslavsky, “Emotional disturbances after stroke.” Clinical and Experimental Hypertension, vol. 28, no. 3-4, pp. 243–249, 2006.

[65] B. Mirheidari, S. M. Bell, K. Harkness, D. Blackburn, and H. Christensen, “Spoken language-based automatic cognitive assessment of stroke survivors.” Language and Health, vol. 2, no. 1, pp. 32–38, 2024.

